# X-chromosome methylation patterns in females with multiple sclerosis

**DOI:** 10.64898/2026.10.13.26365261

**Authors:** Thomas Calnin, Vicki E. Maltby, Sean M. Burnard, Alexandre Xavier, Steve Simpson-Yap, Ingrid van der Mei, Simon Broadley, Bruce V. Taylor, Helmut Butzkueven, Anne-Louise Ponsonby, Rodney J. Scott, Maja Jagodic, Vilija G. Jokubaitis, Jeannette Lechner-Scott, Rodney A. Lea

## Abstract

Multiple sclerosis (MS) is approximately three times more common in females than in males, but sex-linked epigenetic mechanisms remain incompletely understood. We tested whether X-chromosome DNA methylation differs in females with MS and whether associated sites preferentially map to genes normally subject to X-chromosome inactivation (XCI). An X-chromosome-focused epigenome-wide association study was performed in whole blood from three datasets comprising 496 females with MS and 509 female controls, with independent evaluation in isolated CD4+ T cells (80 cases, 76 controls). At FDR < 0.01, 3 182 X-chromosome CpGs were hypomethylated in MS (mean Δβ = −0.035). Hypomethylated CpGs were strongly enriched in non-escape genes normally subject to XCI relative to escape genes (OR 6.77; p = 1.55×10) and mapped to immune and chromatin-related processes. Exploratory cis-mQTL analysis identified strong local genetic effects at a small subset of loci, including WAS, IRAK1 and TLR7. In CD4+ T cells, 2 080 of the 3 182 whole-blood hypomethylated CpGs (65.4%) were also significantly hypomethylated. These findings identify a reproducible X-chromosome hypomethylation signature in female MS and support direct investigation of inactive-X regulation, XCI escape and X-linked immune gene dosage.

## Introduction

Multiple sclerosis (MS) is a chronic immune-mediated and neurodegenerative disorder of the central nervous system with a marked female predominance, with females affected approximately three times more often than males. This pattern has prompted interest in sex-related biological mechanisms, including hormonal, genetic and epigenetic contributors to disease susceptibility. MS is influenced by both genetic susceptibility and environmental exposures, and epigenetic variation, particularly DNA methylation, is a plausible interface between these influences. Changes in DNA methylation have been repeatedly implicated in MS, including at the HLA class II locus and across multiple immune cell types [1–4,14,24]. The X chromosome is a compelling candidate for explaining female-biased autoimmunity. Females undergo dosage compensation through X-chromosome inactivation (XCI), in which one X chromosome is transcriptionally silenced through coordinated chromatin mechanisms, non-coding RNA activity and DNA methylation across the inactive X. XCI is incomplete, and a subset of genes can escape silencing. Here, genes reported to escape XCI are referred to as escape genes, whereas genes normally silenced on the inactive X are referred to as non-escape genes [15]. Variation in XCI escape across tissues and immune states can increase the functional dosage of immune-relevant X-linked genes, such as *TLR7*, with downstream effects on immune activation and signalling [5,6].

Evidence linking XCI dysregulation to female-biased autoimmunity is strongest in systemic lupus erythematosus (SLE), where X-linked gene dosage and loss of epigenetic silencing on the inactive X in T cells have been reported. Demethylation of *CD40LG* on the inactive X has been linked to increased expression in T cells, providing a precedent for autoimmune disease associated with impaired inactive-X silencing [8–10]. *XIST*, a long non-coding RNA expressed from the inactive X chromosome, is central to the establishment and maintenance of XCI, and *XIST*-associated chromatin regulation has been implicated in immune-cell control of inactive-X silencing [6,11,12].

Dedicated XCI-framed analyses of X-chromosome methylation in MS remain limited despite growing evidence for methylation changes in MS blood and immune cells [14,24]. We therefore profiled X-chromosome DNA methylation in females with MS and female controls using array-based whole-blood data and evaluated significant findings in isolated female CD4+ T cells. The aims were to identify X-chromosome differentially methylated positions (DMPs) associated with MS in females, test whether MS-associated DMPs are enriched in genes normally silenced through XCI, and determine whether the findings are reproduced in an independent immune-cell dataset.

## Materials and methods

### Study design and setting

This was an X-chromosome-focused observational case-control epigenome-wide association study (EWAS) using existing whole-blood datasets from Australia and Sweden, with independent evaluation in isolated female CD4+ T cells.

### Participants and data sources

Whole-blood data were obtained from three pre-existing datasets analysed jointly: the Ausimmune Study (208 participants recruited following a first clinical diagnosis of central nervous system demyelination; 402 controls), the Epidemiological Investigation of Multiple Sclerosis (EIMS; 140 MS cases; 139 healthy controls), and an Australian severity dataset (235 female participants with relapse-onset MS) with 102 publicly sourced non-MS control samples [2,14,22,23].

The datasets differed in disease stage, treatment exposure, platform and control source. Ausimmune participants were recruited in four Australian regions during 2003–2006 at a first clinical diagnosis of central nervous system demyelination, with population controls matched at recruitment on age, sex and study region [22]. EIMS samples were collected in Sweden during 2005–2011; 65% of cases were receiving treatment at sampling, predominantly low-efficacy therapies [2]. The Australian severity dataset comprised 235 females with relapse-onset MS from specialist centres in Melbourne, Newcastle, Hobart and Adelaide; 64.7% were receiving disease-modifying therapy at blood collection in the source cohort [23]. Because this cohort lacked internal controls, 102 whole-blood EPIC/850K control IDAT files were selected through the EWAS Data Hub using prespecified criteria for age (23–73 years), female sex, platform, tissue and control status [14]. Dataset subgroup was included as a model covariate.

The sex-stratified whole-blood analysis included 496 female cases and 509 female controls, and 87 male cases and 131 male controls (Table 1; Supplementary Table S1). An additional independent dataset comprised isolated female CD4+ T cells from 80 MS cases and 76 controls, profiled on Illumina HumanMethylation450 arrays [21].

### Methylation and genotype data generation

Whole-blood methylation data were generated using Illumina EPIC arrays (Ausimmune and severity datasets) and HumanMethylation450 arrays (EIMS). For the exploratory cis-methylation quantitative trait locus (cis-mQTL) analysis, genotype data were available for 604 Ausimmune participants (464 females and 140 males) and were generated using the Illumina Global Screening Array v2.0 [14].

### Preprocessing, quality control and normalisation

The source MS EWAS data were preprocessed using methods described previously [14]. Briefly, the ChAMP pipeline [16] was used to parse and filter IDAT files, apply beta-mixture quantile (BMIQ) normalisation [17], and perform ComBat batch correction [18], while retaining probes on sex chromosomes for downstream X-chromosome analyses. The final post-quality-control analysis sets comprised 10 220 X-chromosome and 441 863 autosomal CpGs in whole blood, and 8 585 X-chromosome and 349 084 autosomal CpGs in CD4+ T cells.

### Cell-type proportion estimation

Whole-blood analyses adjusted for estimated immune cell-type proportions using reference-based immune cell deconvolution implemented in the EpiDISH R package [19]. Estimated proportions for B cells, natural killer cells, CD4+ T cells, CD8+ T cells, monocytes, neutrophils and eosinophils were included as covariates in the regression models [14].

### Statistical analysis

The primary EWAS tested for X-chromosome DMPs associated with MS in females. Logistic regression used MS status as the outcome and CpG methylation as the main predictor, adjusting for age, estimated cell-type proportions and dataset subgroup. Statistical significance was controlled using the Benjamini–Hochberg false discovery rate (FDR) at 0.01. Effect size (Δβ) was defined as the unadjusted mean methylation β value in cases minus that in controls and was reported separately from the covariate-adjusted logistic-regression p value used for significance testing. Secondary analyses examined directionality across autosomes and the X chromosome, genomic annotation patterns, enrichment in non-escape genes, *XIST* methylation, consistency in isolated CD4+ T cells and exploratory cis SNP–CpG associations.

Escape and non-escape gene categories were assigned using published human XCI escape annotations, recognising that escape status can vary across tissues, individuals and cell states [15].

### Genetic influence on hypomethylated X-chromosome CpGs

We performed an exploratory cis SNP–CpG association analysis restricted to the 3 182 X-chromosome CpGs hypomethylated in females with MS at FDR < 0.01. CpG and SNP coordinates were based on hg19. Directly genotyped X-chromosome SNPs from the Illumina Global Screening Array v2.0 within ±2 000 bp of each CpG were identified. Genotypes were coded additively as dosage values of 0, 1 or 2 and matched to methylation profiles by participant identifier. The final matched female Ausimmune dataset included 464 individuals. Because CpGs were selected on MS-associated hypomethylation but SNP–CpG associations were tested in cases and controls combined, this analysis was interpreted as a cis-mQTL screen rather than a test of disease-specific genetic effects.

For each cis SNP–CpG pair, methylation β value was modelled as a function of SNP dosage using unadjusted linear regression. Associations were excluded where fewer than 20 matched samples or fewer than two genotype dosage classes were available. After filtering, 2 312 SNP– CpG pairs were tested, representing 1 469 CpGs and 593 SNPs, with complete-case sample sizes of 444–464. The SNP-dosage p value was assessed against a conservative threshold of p < 5×10 ; among significant associations, strength was summarised using r².

### Functional enrichment

Gene Ontology (GO) biological process over-representation analysis was performed in R using 623 unique genes annotated to hypomethylated X-chromosome CpGs identified in females with MS at FDR < 0.01. Analysis used clusterProfiler v4.14.6 [20], with visualisation using enrichplot. The primary analysis used FDR correction and an X-chromosome-only background defined by genes represented by probes detected on the array, passing quality control and mapping to the X chromosome.

## Results

### Participants and analysis sets

The pooled whole-blood EWAS comprised the Ausimmune, EIMS and Australian severity/public-control datasets summarised in Figure 1. The pooled analysis comprised three constituent datasets rather than separate discovery and validation stages. Participant characteristics are shown in Table 1 and Supplementary Table S1.

**Figure 1.**
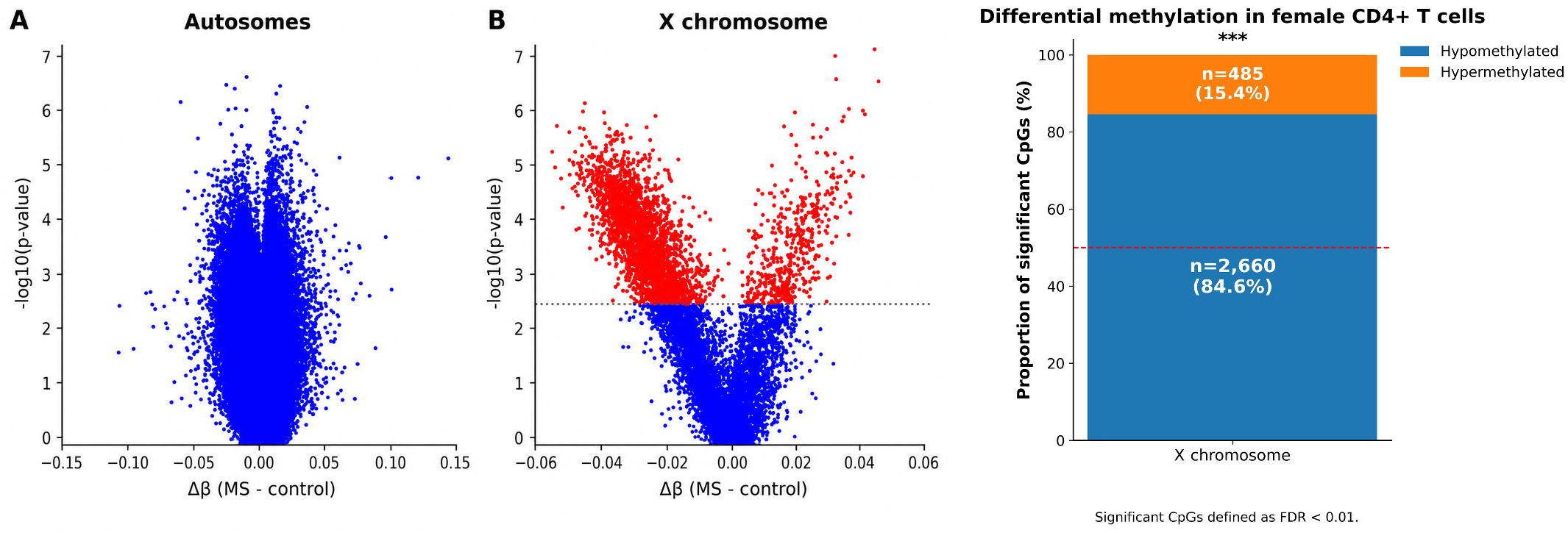
Study datasets, biospecimens and analysis overview. Schematic of the three whole-blood datasets, sex-stratified EWAS analysis sets, the female Ausimmune cis-mQTL subset and isolated female CD4+ T-cell validation, including array platforms and high-level analysis steps.

### Female MS is associated with predominantly hypomethylated X-chromosome DMPs

In females, the X chromosome showed widespread hypomethylation in MS compared with controls, with 3 182 significantly hypomethylated X-chromosome CpGs at FDR < 0.01. No CpGs reached FDR < 0.01 on either the X chromosome or autosomes in the smaller male analysis (Figure 2).

**Figure 2.**
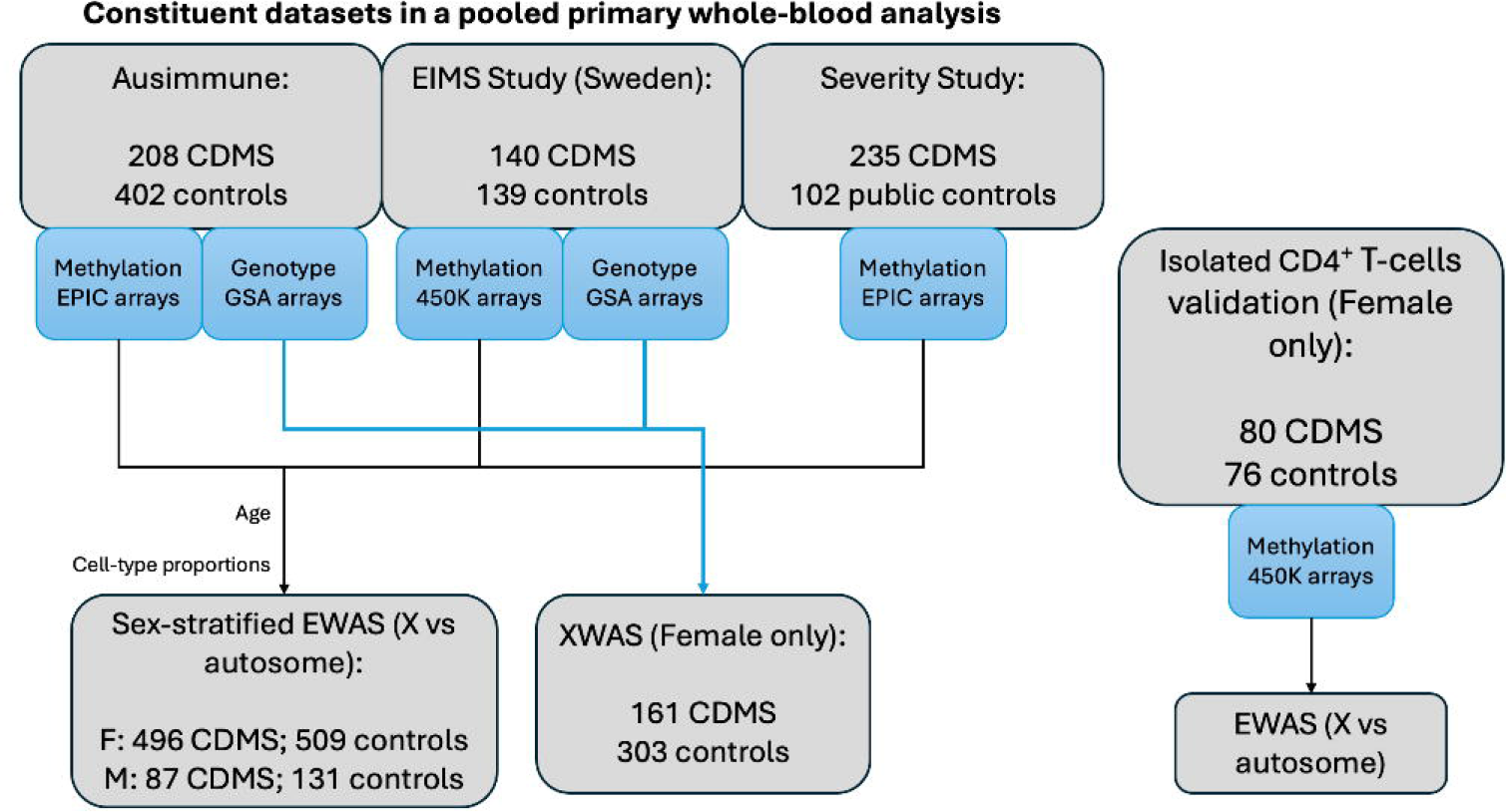
Sex-stratified EWAS results on autosomes and the X chromosome. Volcano plots show results for female autosomes, the female X chromosome, male autosomes and the male X chromosome. The x-axis represents mean Δβ (mean βMS − mean βcontrol) and the y-axis represents −log10 nominal p value from the covariate-adjusted logistic-regression model. Summary bars depict the directionality of significant CpGs in females. Significant CpGs were defined at FDR < 0.01; no CpGs reached this threshold in either male analysis.

Across chromosomes in females, significant autosomal CpGs were predominantly hypermethylated (18 384 hypermethylated versus 5 799 hypomethylated), whereas significant X-chromosome CpGs were predominantly hypomethylated (3 182 hypomethylated versus 432 hypermethylated) (Figure 2; Supplementary Figure S5).

### Genomic annotation and representative loci

Effect sizes among the 3 182 hypomethylated X-chromosome CpGs were modest but consistent (mean Δβ = −0.035; median Δβ = −0.035; range −0.064 to −0.0008). Most annotated CpGs mapped to CpG islands or shores, and promoter-proximal annotations were common (Supplementary Figures S1 and S2). Gene-symbol annotation was unavailable for 329 probes (10.3%). Recurrently represented loci included *BCYRN1*, *FGF13*, *TIMM17B*, *PQBP1*, *NHSL2*, *ARX*, *WDR45*, *TSC22D3*, *IKBKG* and *G6PD*.

Among the most statistically significant probes was cg07759587, annotated to *TLR8*/LOC349408 (Δβ = −0.030; FDR = 2.88×10), with other highly significant probes mapping to *ACSL4*, *XIAP*, *BEX1* and *DOCK11*. The largest methylation differences included cg18717600, annotated to *ZNF674*/LOC401588 (Δβ = −0.064; FDR = 4.11×10), and probes mapping to *PHF16*, *MAP7D3* and *RAP2C*. Representative DMPs are shown in Table 2.

### Enrichment in X-inactivation genes

To assess whether hypomethylation preferentially involved genes normally subject to XCI, significant CpGs were compared across escape and non-escape categories. Hypomethylated CpGs were over-represented in non-escape genes relative to escape genes (OR 6.77; p = 1.55×10 ; Table 3). XIST methylation pattern At assayed CpGs mapping to *XIST* (cg03554089, cg05533223, cg17279685, cg11717280, cg02644889, cg17513789, cg20698282 and cg12653510), mean methylation differed between MS cases and controls, with several probes showing small but consistent hypermethylation in cases (Figure 3). These site-specific differences do not establish the direction of *XIST* expression or inactive-X maintenance.

**Figure 3.**
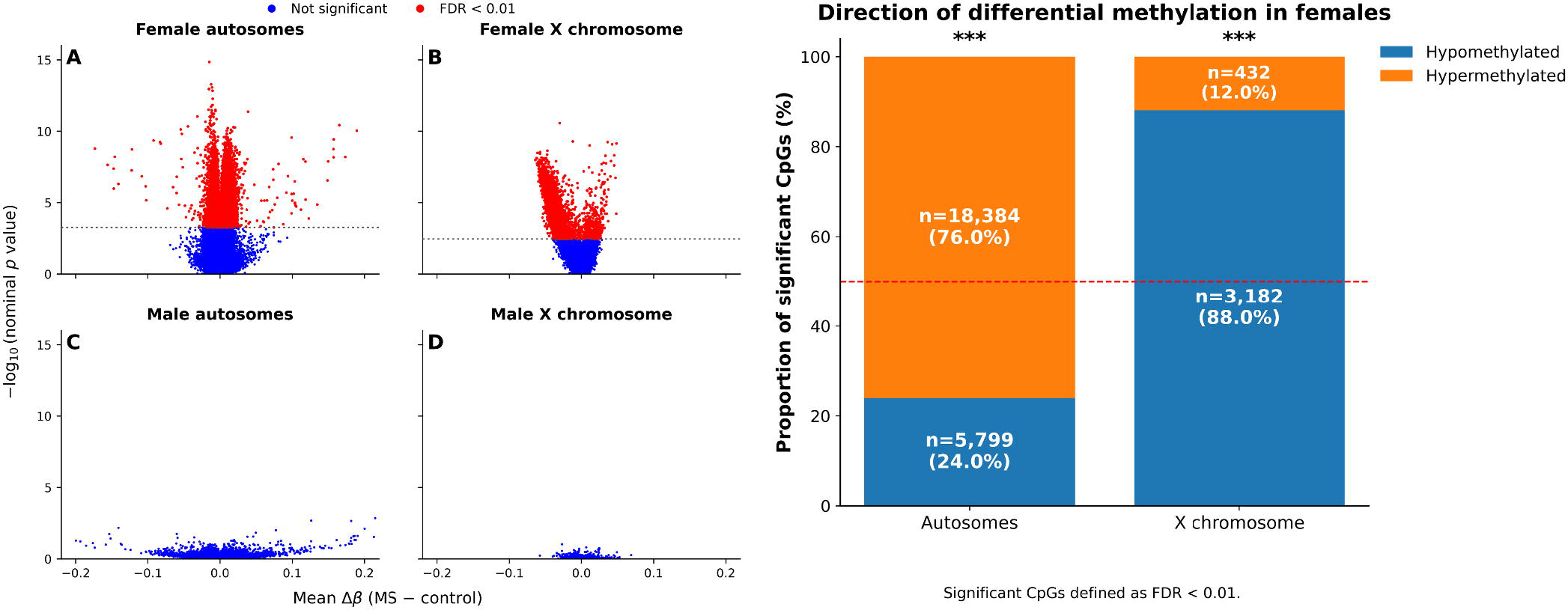
XIST methylation profile in females with multiple sclerosis. Mean methylation β values at assayed *XIST* CpG sites in females with MS and female controls. Asterisks indicate nominally significant case-control differences (*p < 0.05; **p < 0.01; ***p < 0.001).

### Functional enrichment

GO biological process analysis identified 167 terms significant after FDR correction using an X-chromosome-only background. Immune system process was over-represented (61 genes; fold enrichment 1.33; adjusted p = 8.99×10), together with chromatin organisation (42 genes; fold enrichment 1.41; adjusted p = 1.13×10 ³) and chromatin remodelling (34 genes; fold enrichment 1.45; adjusted p = 1.61×10 ³). Additional immune-related terms included regulation of immune system process, immune response and innate immune response (Supplementary Figure S3).

### Genetic influence on hypomethylated CpGs

Among 2 312 tested cis SNP–CpG pairs, 28 reached p < 5×10, involving 16 unique CpGs and 28 unique SNPs. Significant CpGs mapped to *WAS*, *IRAK1*, *TLR7*, *PRKX*, *SHROOM2*, *PRDX4*, *HTR2C*, *CXorf22*, *ABCB7*, *GPR173*, *STARD8*, *MBTPS2*, *SLITRK4* and *NKRF*, with two significant CpGs mapping to *HTR2C* and one lacking gene-symbol annotation.

The strongest association by r² was rs145299197–cg24428913 at *WAS* (β = −0.318, p = 9.77×10 ¹, r² = 0.769) (Figure 4). Other strong associations included rs1059702– cg02742918 at *IRAK1* (p = 2.55×10 ³, r² = 0.297), rs2302267–cg24735671 at *TLR7* (p = 3.49×10 ², r² = 0.210), rs1003351–cg22364675 at *PRKX* (p = 5.23×10 ²¹, r² = 0.175) and rs3788949–cg13185972 at *SHROOM2* (p = 3.12×10 ¹, r² = 0.126). Most hypomethylated CpGs did not show a significant local cis association within the tested ±2 000-bp window.

**Figure 4.**
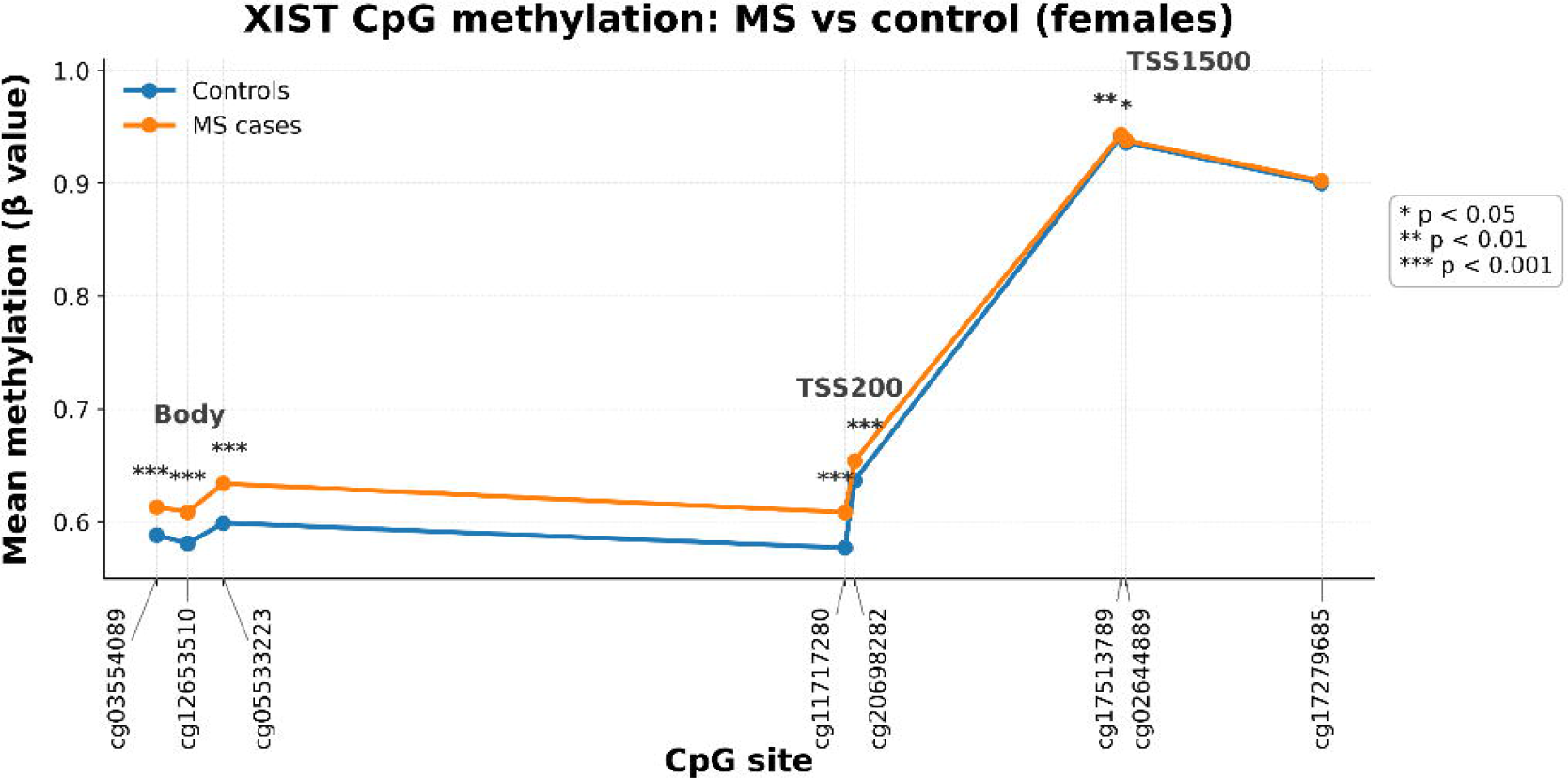
cis SNP–CpG associations for hypomethylated X-chromosome CpGs. Manhattan-style plot of cis SNP–CpG associations for hypomethylated X-chromosome CpGs in females. Directly genotyped SNPs within ±2 000 bp of each CpG were tested against methylation β value using additive SNP dosage. The y-axis shows −log10(p) for the SNP-dosage term; the dashed line denotes p = 5×10 . The largest outlying point corresponds to rs145299197–cg24428913 (*WAS*).

### Independent CD4+ T-cell data replicate X-chromosome hypomethylation

In isolated female CD4+ T cells (80 MS cases; 76 controls), 3 145 X-chromosome DMPs reached FDR < 0.01, of which 2 660 were hypomethylated and 485 were hypermethylated.

Effect sizes were modest overall (mean Δβ = −0.021; median Δβ = −0.026; range −0.055 to 0.042), and 2 204 CpGs were significant in both whole blood and CD4+ T cells. Of the 3 182 whole-blood hypomethylated X-chromosome DMPs, 2 080 (65.4%) were also significant and hypomethylated in CD4+ T cells, providing substantial independent replication of the whole-blood signal (Figure 5).

**Figure 5.**
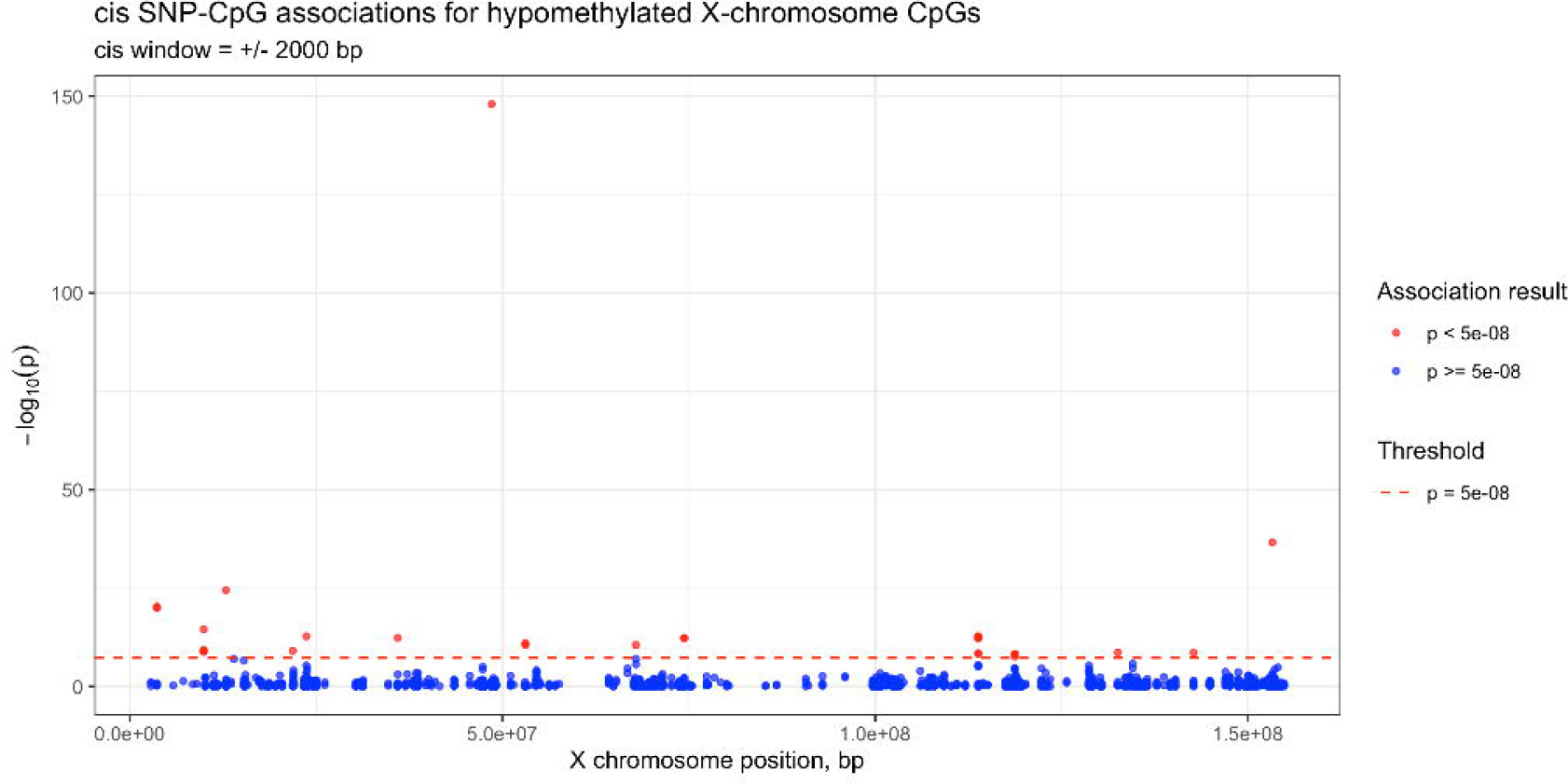
Independent CD4+ T-cell validation. Volcano plots and directionality summary for isolated female CD4+ T cells (80 MS cases; 76 controls), demonstrating predominant X-chromosome hypomethylation compared with autosomes. Significant CpGs were defined at FDR < 0.01.

## Discussion

Across three whole-blood datasets, females with MS showed a strongly directional pattern of X-chromosome methylation change characterised by widespread hypomethylation: 3 182 X-chromosome CpGs were significantly hypomethylated at FDR < 0.01, with modest but consistent effect sizes. No CpGs reached FDR < 0.01 in the smaller male analysis; this lack of significance should not be interpreted as evidence that the pattern is female-specific.

A key finding was enrichment of hypomethylated CpGs in non-escape genes, that is, genes usually silenced on the inactive X (Table 3). Considered alongside case-control methylation differences at *XIST* probes (Figure 3), this pattern is compatible with altered regulation or maintenance of XCI in females with MS, although the data do not establish mechanism.

Importantly, 2 080 of the 3 182 whole-blood hypomethylated DMPs (65.4%) were also significant and hypomethylated in isolated female CD4+ T cells (Figure 5), supporting an immune-cell-intrinsic component beyond whole-blood cell-mixture effects.

Sex-biased autoimmunity has long raised the possibility that X-linked gene dosage and XCI escape contribute to female risk. Prior work in SLE supports loss of epigenetic silencing on the inactive X in T cells, including *CD40LG* demethylation and X-linked gene overexpression, and *TLR7* can escape XCI in immune cells with downstream interferon consequences [5,8–10].

Absher et al. reported a broader pattern of X-chromosome hypomethylation in CD4+ T cells in SLE [13], providing disease-contextual support for an X-linked epigenetic axis in female-predominant autoimmunity.

The present findings extend this concept to MS. If hypomethylation preferentially affects genes normally silenced on the inactive X, partial relaxation of XCI maintenance in specific immune lineages is one plausible mechanism by which functional dosage of X-linked immune genes could increase. This remains hypothesis-generating (Supplementary Figure S4): methylation at the assayed sites does not directly measure XCI escape or transcription, and the *XIST* methylation pattern requires locus-specific interpretation and direct transcript quantification. The cis-mQTL analysis identified strong local genetic effects at a small subset of DMPs, including immune-relevant loci such as *WAS*, *IRAK1* and *TLR7*, but did not account for most of the signature within the tested window.

Strengths of the study include explicit X-chromosome analysis within a sex-stratified framework, integration of multiple whole-blood datasets, stringent FDR control, and independent evaluation in purified CD4+ T cells. Several limitations should also be considered. Residual confounding by cell mixture, dataset heterogeneity, smoking, treatment exposure, disease duration, phenotype and other incompletely harmonised factors cannot be excluded. Public controls used with the severity cohort differed from cases in source and age distribution, and covariate adjustment may not fully remove this bias. Methylation changes do not necessarily imply altered expression, and inference regarding XCI maintenance or escape requires transcriptomic and allele-specific confirmation. The male analysis was underpowered relative to the female analysis.

In summary, females with MS show widespread, modest X-chromosome hypomethylation enriched in genes normally subject to XCI, with substantial replication in CD4+ T cells. Future work should combine methylation with gene-expression and allele-specific analyses across relevant immune subsets to determine whether the observed signature reflects altered XCI maintenance, XCI escape or X-linked immune gene dosage, and should examine age-related, environmental and reproductive modifiers of this signal.

## Data Availability

The EIMS whole-blood DNA methylation data are publicly available through the NCBI Gene Expression Omnibus (GEO; accession GSE106648; https://www.ncbi.nlm.nih.gov/geo/query/acc.cgi?acc=GSE106648). Individual-level methylation, genotype and linked clinical data from the remaining cohorts are not publicly available because of restrictions associated with participant consent, research ethics approvals and cohort data-governance agreements. These restrictions prevent unrestricted dissemination of participant-level genomic and clinical information.

## Acknowledgements

The authors acknowledge the Ausimmune Investigator team, including Robyn Lucas, Patricia Valery, Keith Dear and Caron Chapman, and thank all participants and research personnel who contributed to the Ausimmune and AusLong studies. The authors also acknowledge Susan Agland, Barbara Alexander, Marcia Davis, Zoe Dunlop, Rosalie Scott, Marie Steele, Catherine Turner, Brenda Wood, Jane Gresham, Camilla Jozwick, Helen Rodgers, Carol Hurst, Kate Smith, Talia Nichols, Claire Knowles and Roxanne Maher for study coordination, recruitment, data collection and follow-up activities.

## Ethics statement

The Ausimmune Study was approved by nine regional Human Research Ethics Committees (study number 09/04/15/5.04 and HREC/09/HNE/139). The CD4+ T-cell and severity studies were approved by the Hunter New England Human Research Ethics Committee (2019/ETH12346). EIMS was approved by the Regional Ethical Review Board at Karolinska Institutet, Stockholm, Sweden (2004-252/1-4). All participants provided written informed consent.

## Author contributions

TC contributed to data analysis, visualisation and manuscript drafting. VEM contributed to study conception, cohort and data acquisition, interpretation, supervision and manuscript revision. SMB contributed to data curation, methylation and genotype processing, analysis and manuscript revision. AX contributed to methodology, bioinformatic analysis, data curation and manuscript revision. ALP and BVT contributed to Ausimmune/AusLong study design, cohort resources, data acquisition and manuscript revision. The Ausimmune/AusLong Investigators contributed to participant recruitment, study conduct, data collection and cohort resources. RJS contributed to resources, data generation, supervision and manuscript revision. MJ contributed EIMS resources and epigenetic expertise, interpretation and manuscript revision. VGJ contributed severity-cohort resources, interpretation and manuscript revision. JLS contributed clinical cohort leadership, resources, supervision, interpretation and manuscript revision. RAL conceived and supervised the X-chromosome analysis, contributed methodology and interpretation, and led manuscript drafting and revision. All authors reviewed and approved the final manuscript.

## Use of generative AI

ChatGPT (OpenAI, San Francisco, CA, USA) was used to assist with language editing and journal-specific manuscript formatting. It was not used to generate or analyse study data. All scientific content, analyses, interpretations and final text were reviewed and approved by the authors.

