## Supplementary Figures for "X-chromosome methylation patterns in females with multiple sclerosis"

#### Contents

Supplementary Figure S1. Annotation summary of hypomethylated X-chromosome CpGs

Supplementary Figure S2. Gene-region distribution of hypomethylated X-chromosome CpGs

Supplementary Figure S3. Gene Ontology biological process over-representation analysis

Supplementary Figure S4. Conceptual model of XCI dysregulation in multiple sclerosis

Supplementary Figure S5. Female autosomal differential methylation by chromosome

Supplementary Table S1. Baseline characteristics pooled across the three whole-blood datasets

### Supplementary Figure S1. Annotation summary of hypomethylated X-chromosome CpGs.

Summary of effect sizes and genomic context for 3 182 significant hypomethylated X-chromosome CpGs (FDR < 0.01), including CpG island context and recurrently annotated loci.

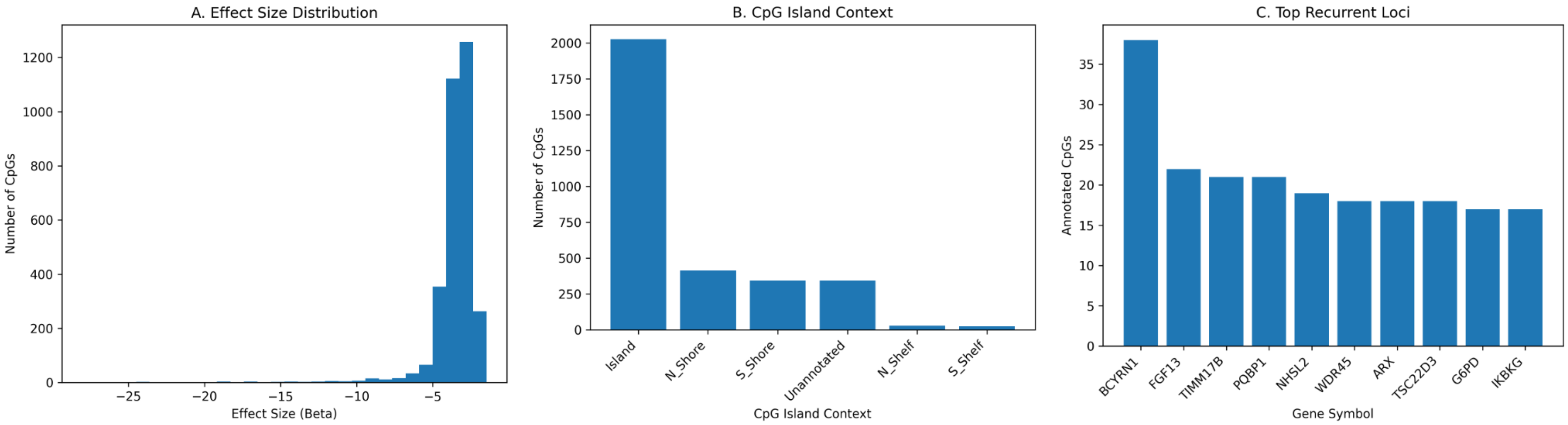

### Supplementary Figure S2. Gene-region distribution of hypomethylated X-chromosome CpGs.

Numbers of significantly hypomethylated CpG probes annotated to RefGene gene-region categories. CpGs with multiple region annotations may contribute to more than one category; unannotated denotes CpGs without a GENE_GROUP assignment.

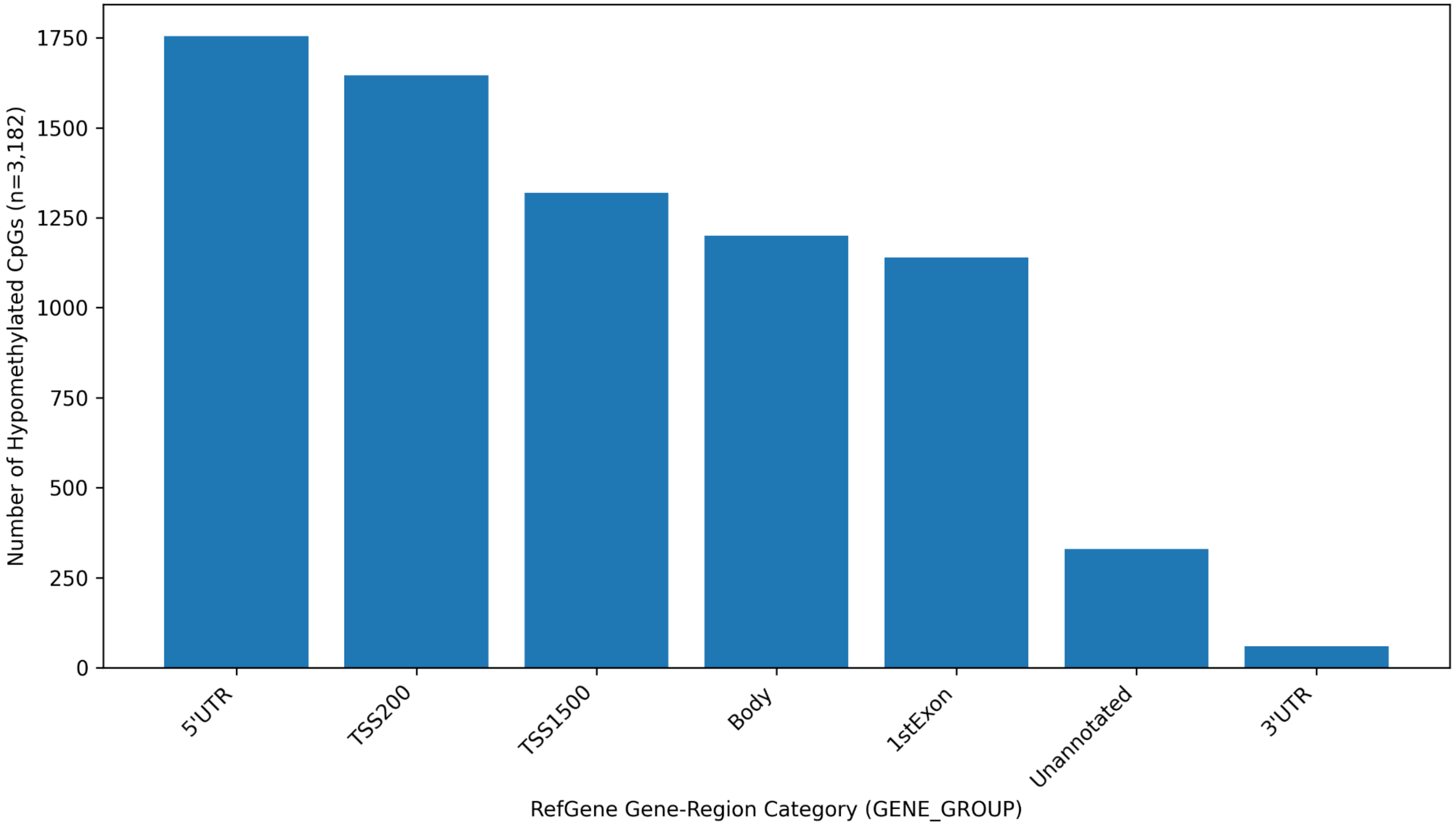

### Supplementary Figure S3. Gene Ontology biological process over-representation analysis.

Tree plot of genes annotated to hypomethylated X-chromosome CpGs using an X-chromosome-only background. Related enriched terms are grouped by pairwise similarity; circle size indicates contributing gene count and colour indicates FDR-adjusted p value.

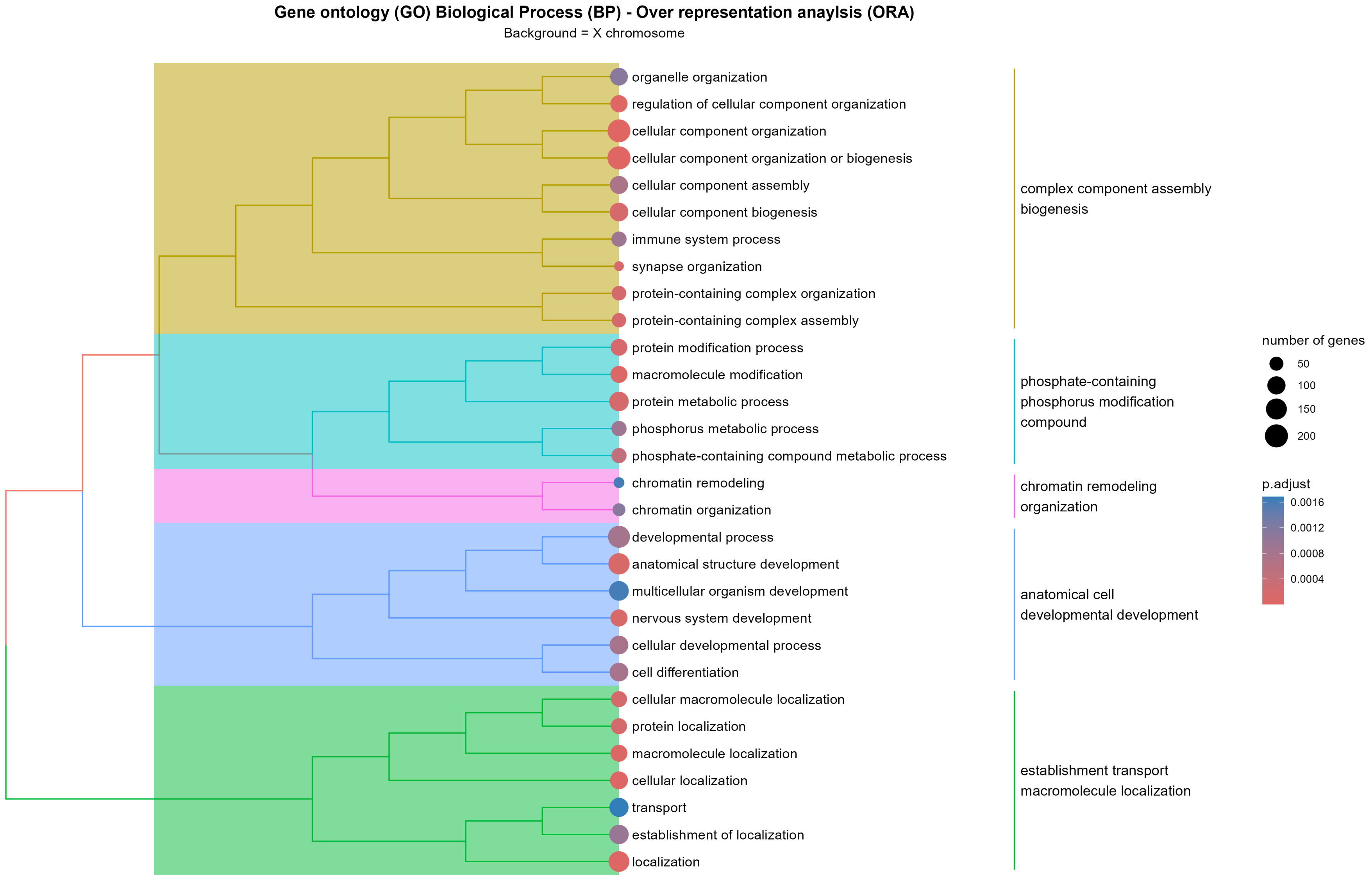

### Supplementary Figure S4. Conceptual model of XCI dysregulation in multiple sclerosis.

Hypothesis-generating schematic linking altered XIST/DNMT-associated regulation to X-chromosome hypomethylation and potential reactivation or dosage effects in immune cells. The model is conceptual and does not constitute direct mechanistic evidence.

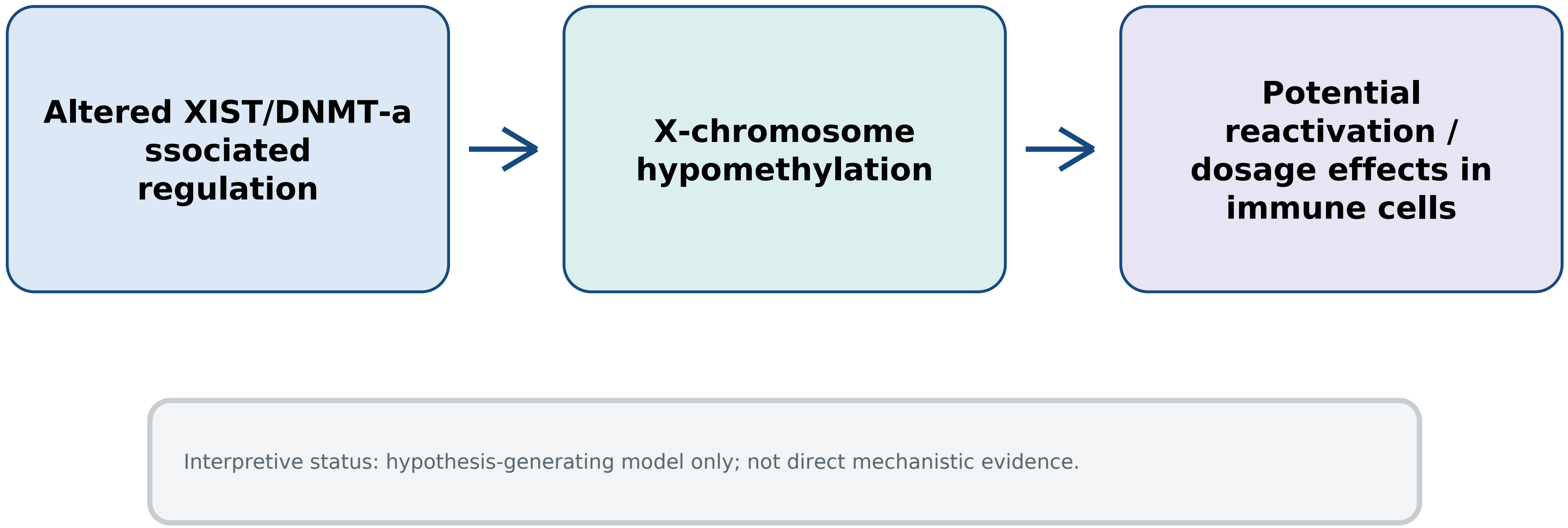

### Supplementary Figure S5. Female autosomal differential methylation by chromosome.

Volcano plots of female MS-associated differential methylation by autosome using FDR < 0.01. Although chromosome-specific patterns vary, significant autosomal CpGs show an overall tendency toward hypermethylation, in contrast to the predominant X-chromosome hypomethylation.

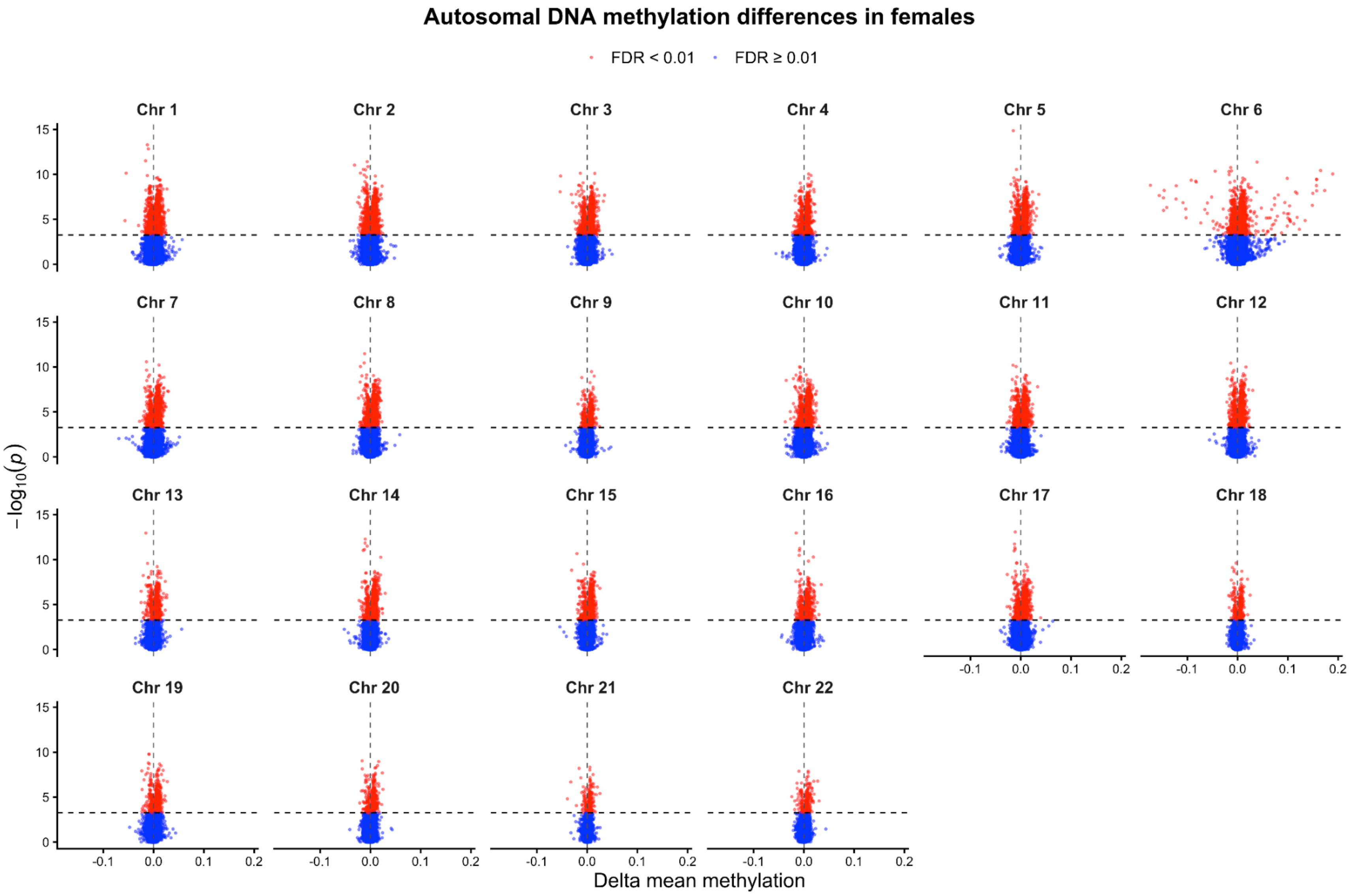

### Supplementary Table S1. Baseline characteristics pooled across the three whole-blood datasets.

| Characteristic | Female MS (n = 496) | Female controls (n = 509) | Male MS (n = 87) | Male controls (n = 131) |
|---|---|---|---|---|
| **Age, years, mean (SD)** | 43.8 (11.4) | 44.3 (12.2) | 39.1 (11.0) | 40.8 (10.3) |
| **Treatment status at sampling, n (%)** | IFN-β: 55 (11.1%)<br>None: 218 (44.0%)<br>Missing/other: 223 (45.0%) | NA | IFN-β: 8 (9.2%)<br>None: 36 (41.4%)<br>Missing/other: 43 (49.4%) | NA |
| **Estimated cell proportions (summary)** | B: 0.047<br>Mono: 0.055<br>NK: 0.036<br>CD8T: 0.092<br>CD4T: 0.130<br>Neutro: 0.616<br>Eos: 0.007 | B: 0.044<br>Mono: 0.050<br>NK: 0.043<br>CD8T: 0.105<br>CD4T: 0.121<br>Neutro: 0.609<br>Eos: 0.008 | B: 0.052<br>Mono: 0.065<br>NK: 0.043<br>CD8T: 0.088<br>CD4T: 0.121<br>Neutro: 0.601<br>Eos: 0.009 | B: 0.046<br>Mono: 0.066<br>NK: 0.048<br>CD8T: 0.098<br>CD4T: 0.115<br>Neutro: 0.594<br>Eos: 0.012 |
| **Dataset membership (Ausimmune/EIMS/Severity), n** | 163 / 98 / 235 | 303 / 104 / 102 | 45 / 42 / 0 | 96 / 35 / 0 |

Note. Detailed disease duration, phenotype and smoking variables were not uniformly harmonised across contributing datasets and were not included as covariates in the primary EWAS; residual confounding cannot be excluded.
